# Targeted Quantitative Plasma Metabolomics Identifies Metabolite Signatures that Distinguish Heart Failure with Reduced and Preserved Ejection Fraction

**DOI:** 10.1101/2024.07.24.24310961

**Authors:** Fawaz Naeem, Teresa C. Leone, Christopher Petucci, Clarissa Shoffler, Ravindra C. Kodihalli, Tiffany Hidalgo, Cheryl Tow-Keogh, Jessica Mancuso, Iphigenia Tzameli, Donald Bennett, John D. Groarke, Rachel J. Roth Flach, Daniel J. Rader, Daniel P. Kelly

**Author notes:** Corresponding Author: Daniel P. Kelly, M.D. Perelman School of Medicine, University of Pennsylvania 3400 Civic Center Blvd. Smilow Translational Research Center, Room 11-122 Philadelphia, PA 19104.

## Abstract

**Background:** Two general phenotypes of heart failure (HF) are recognized: HF with reduced ejection fraction (HFrEF) and with preserved EF (HFpEF). To develop HF disease phenotype-specific approaches to define and guide treatment, distinguishing biomarkers are needed. The goal of this study was to utilize quantitative metabolomics on a large, diverse population to replicate and extend existing knowledge of the plasma metabolic signatures in human HF.

**Methods:** Quantitative, targeted LC/MS plasma metabolomics was conducted on 787 samples collected by the Penn Medicine BioBank from subjects with HFrEF (n=219), HFpEF (n=357), and matched non-failing Controls (n=211). A total of 90 metabolites were analyzed, comprising 28 amino acids, 8 organic acids, and 54 acylcarnitines. 733 of these samples were also processed via an OLINK protein panel for proteomic profiling.

**Results:** Consistent with previous studies, unsaturated forms of medium/long chain acylcarnitines were elevated in the HFrEF group to a greater extent than the HFpEF group compared to Controls. A number of amino acid derivatives, including 1- and 3-methylhistidine, homocitrulline, and symmetric (SDMA) and asymmetric (ADMA) dimethylarginine were elevated in HF, with ADMA elevated uniquely in HFpEF. Plasma branched-chain amino acids (BCAA) were not different across the groups; however, short-chain acylcarnitine species indicative of BCAA catabolism were significantly elevated in both HF groups. The ketone body 3-hydroxybutyrate (3-HBA) and its metabolite C4-OH carnitine were uniquely elevated in the HFrEF group. Linear regression models demonstrated a significant correlation between plasma 3-HBA and NT-proBNP in both forms of HF, stronger in HFrEF.

**Conclusions:** These results identify plasma signatures that are shared as well as potentially distinguish between HFrEF and HFpEF. Metabolite markers for ketogenic metabolic re-programming in extra-cardiac tissues were identified as unique signatures in the HFrEF group, possibly related to the lipolytic action of increased levels of BNP. Future studies will be necessary to further validate these metabolites as HF biosignatures that may guide phenotype-specific therapeutics and provide insight into the systemic metabolic responses to HFpEF and HFrEF.

**Clinical Perspective:** *What Is New?:*

- “Real world” targeted metabolomic profiling on wide range of metabolites in a diverse population of patients with HFrEF and HFpEF.
- Levels of 3-hydroxybutyrate and its metabolite C4OH-carnitine were uniquely increased in the HFrEF group and correlated with levels of plasma NT-proBNP in both the heart failure groups, indicating the possibility of a heart-adipose-liver axis.
- Asymmetric dimethylarginine, a known inhibitor of nitric oxide synthase, was uniquely upregulated in HFpEF suggesting that there may also be an underlying component of vascular dysregulation contributing to HFpEF pathophysiology.

*What Are the Clinical Implications?:*

- The plasma metabolomic changes seen in the heart failure cohorts support the existing theory of metabolic reprogramming, providing further rationale for the pursuit of therapeutic targets for the treatment of heart failure.
- Quantitative metabolomic profiling shows promise for guiding therapeutic decisions in HFrEF and HFpEF.
- Modulation of natriuretic peptides may enhance the delivery of ketone and fatty acids to the “fuel starved” failing heart.

## Introduction

Despite recent advances in treatment, heart failure (HF) remains a major cause of cardiovascular mortality worldwide and has a profound impact on functionality and quality of life.^1^ HF is a broad clinical syndrome encompassing a heterogeneous group of diseases defined, in part, by degree of ventricular functional impairment and remodeling. While there is some debate regarding classification, two predominant phenotypes are recognized: HF with reduced ejection fraction (HFrEF) and HF with preserved ejection fraction (HFpEF).^2^ Our understanding of the pathogenic drivers that distinguish among these HF groups is incomplete, and therefore treatment regimens are generally not tailored to HF phenotype. Accordingly, identification of specific biosignatures to delineate distinct HF phenotypes is an important unmet need and could ultimately assist in the development of disease phenotype-targeted therapeutics.

Unbiased transcriptomic, metabolomic and/or proteomic profiling of myocardial tissues have been conducted in preclinical (rodent) models of HFrEF.^3–6^ The results of these metabolomic profiling studies are consistent with the known shifts in myocardial fuel utilization that occur in HFrEF, including reduced capacity for mitochondrial fatty acid oxidation (FAO)^7–13^ as reflected, in part, by reduced levels of ventricular medium- (MCAC) and long-chain acylcarnitines (LCAC), intermediates generated in the mitochondrial β-oxidation of fatty acids (FAs). Cardiac metabolomic analyses in pre-clinical models of HF have also identified signatures of decreased catabolism of branched-chain amino acids (BCAAs)^3,14,15^ and increased oxidation of ketone bodies^5^ in HFrEF myocardium, the latter likely used as an ancillary fuel in the context of diminished utilization of FA, the chief fuel of the normal adult heart. A small number of myocardial tissue-based metabolomic analyses in humans with HFrEF have also been reported.^16–18^ The results of these studies are consistent with the pre-clinical studies demonstrating reduced MCAC/LCAC and shorter-chain ACs indicative of reduced BCAA degradation. In addition, one study using endomyocardial biopsies in HFpEF patients and post-transplant or donor heart tissue in HFrEF patients demonstrated signatures of reduced MCAC/LCAC in both HFrEF and HFpEF along with increased levels of the ketone body, 3-hydroxybutyrate (3-HBA), in the HFrEF myocardium.^18^ Lastly, a recent seminal study in which metabolites were collected from both systemic arterial and coronary sinus in humans to assess myocardial extraction levels found that the FA utilization was decreased and ketone utilization increased in humans with HFrEF compared to normal controls.^19^

A number of studies have assessed plasma metabolomics in humans with HF, particularly HFrEF, with a few studies comparing HFrEF and HFpEF.^18,20–22^ The majority of HFrEF studies have shown an increase in circulating MCAC and LCAC.^18,20,21,23,24^ This is a rather surprising finding given that these metabolites are reduced in HFrEF myocardium, suggesting that the origin of these species are likely extra-cardiac. The limited data available in which the plasma metabolome from HFrEF and HFpEF patients have been compared is conflicting, likely related to phenotypic heterogeneity and relatively low subject numbers.^18,20^ For example, information on plasma levels of BCAA and ketone bodies are inconclusive. Zordoky et al. found that plasma ketones were increased in HFpEF to a greater extent than HFrEF.^21^ A larger study conducted by Hunter et al. found no changes in ketone and ketone-related metabolites in the plasma of HF versus controls.^20^ They also found that BCAAs and their metabolites were uniquely upregulated in HFrEF. In contrast, Hahn et al. recently found that circulating ketone bodies were elevated in HFrEF compared to HFpEF patients, with minimal changes in BCAAs.^18^ In addition to these targeted plasma studies, there have been several studies that have utilized untargeted metabolomics to identify potential biomarker panels that predict HF severity and mortality.^25–27^

Given the relative paucity and conflicting data regarding circulating metabolites in HFrEF versus HFpEF, we sought to conduct a study in a relatively large and diverse population of humans with HFrEF, HFpEF, and corresponding controls without HF in order to replicate and extend existing knowledge regarding potential distinguishing biosignatures. Notably, most of the studies cited above lacked large study populations and racial/ethnic diversity. In this study, we conducted quantitative plasma metabolomics and targeted proteomics on a large, diverse sample of patients. We collected samples in the ad lib fed state with an aim towards identifying markers that show promise for “real world” application. Our results indicate that a significant subset of unsaturated MCAC and LCAC were increased uniquely in the HFrEF group. In addition, circulating metabolites indicative of ketogenesis and ketone oxidation were significantly increased in HFrEF but not HFpEF. Methylated derivatives of arginine were shown to be elevated in HF, with asymmetric dimethylarginine increased uniquely in the HFpEF group. Lastly, BCAA levels were not different among the groups but metabolites indicative of BCAA catabolism were increased in both HFrEF and HFpEF.

## Methods

### Study Population

The present study employed the Penn Medicine Biobank registry to identify anonymized patient data and plasma samples. The Biobank is a research program at the University of Pennsylvania in which enrolled participants provide consent for research, access to their medical records, genetic data, and blood samples. The Biobank was systematically reviewed to identify patients meeting specific inclusion criteria for 1 of 3 groups: HFrEF, HFpEF, or control. Inclusion criteria for the HFrEF group was defined as patients with a clinical diagnosis of HF and an ejection fraction of less than 35%. The HFpEF group consisted of patients with a clinical diagnosis of HF, an ejection fraction greater than 45%, and an H2FPEF score^28^ greater than 4, giving a probability of 70% or higher. Patients with HF secondary to hypertrophic cardiomyopathy, restrictive cardiomyopathy, amyloidosis, or valvular disease were excluded from the study. The control group was generated by matching to the HFpEF group for gender, race, age, and comorbid diagnoses of hypertension and diabetes. Exclusion criteria for the control group included HF of any etiology, cancer diagnosis in the last 5 years, any autoimmune disease, cirrhosis, nonalcoholic steatohepatitis, chronic obstructive pulmonary disease, or idiopathic pulmonary fibrosis.

### Targeted Metabolomics

The Biobank obtained plasma samples of enrolled patients during routine outpatient phlebotomy appointments. Fasting was not a requirement for sampling. Samples were obtained between 2008-2022 and were stored in a -80°C freezer. Following screening of the Biobank and identification of eligible patients, corresponding plasma samples were obtained and sent to the Penn Metabolomics Core for targeted metabolomic analysis. From the plasma samples, aliquots (50-100 µL) were extracted for ACs, amino acids, and organic acids according to validated, optimized protocols as described.^23^ Each class of metabolites was separated with a unique high-performance liquid chromatography (HPLC) method to optimize their chromatographic resolution and sensitivity. Quantitation of metabolites in each assay module was achieved using multiple reaction monitoring of calibration solutions and study samples on an Agilent 1290 Infinity UHPLC/6495B triple quadrupole mass spectrometer. Raw data was processed using Mass Hunter quantitative analysis software (Agilent). Calibration curves (R2 = 0.99 or greater) were either fitted with a linear or a quadratic curve with a 1/X or 1/X2 weighting.

Plasma levels of NT-proBNP were profiled using the Target 96 Cardiovascular III panels (Product #95611A, Lot #B24627A; corresponding detection and controls, Product #95611B, Lot #B24627B) using Olink technologies (www.olink.com; Watertown, MA) by Pfizer, Inc.^29^ This panel allows simultaneous analysis of 92 protein biomarkers with documented or suggested involvement in cardiovascular processes or diseases using 1 µL of sample volume. A total of 733 (eight batches of 88 and one batch of 29) K2 EDTA plasma samples were randomly distributed across 9 plates, and protein analyses were performed by trained technicians blinded to the clinical information. Any failed sample in a given batch or outlier samples were included in the last batch of samples. Each analysis plate, consisting of 96 wells, included two sample controls to assess coefficient precision, three negative controls to establish background levels for each protein assay and calculate the detection limit, and three inter-plate controls to adjust for any variations between runs and plates. The samples were analyzed according to the protocol provided by Olink.

### Statistical Analyses

A total of 90 metabolites were analyzed, comprising 28 amino acids, 8 organic acids, and 54 ACs, which were further classified into short, medium, long, and very-long chain ACs. Metabolites for which greater than 50% of the samples returned above or below the limit of quantification were excluded from the analysis. Statistical comparisons between metabolites and clinical biomarkers were performed using One-Way ANOVA with multiple comparisons. Comorbidity and demographic comparisons were conducted using Fischer’s exact test. Radar plots were generated in Visual Studio using the ChartJS software. Linear regression models with an OLINK Protein Panel, including NTproBNP, as the response variable were used to explore mean differences between HFpEF, HFrEF, and controls as well with covariates including race, sex, age, BMI, MI, ischemia, T2 diabetes, hypertension, hyperlipidemia, atrial fibrillation, stroke, sleep apnea, and chronic kidney disease. Tukey’s adjustment was used for multiple comparisons within an independent variable. Histograms, QQ plots, and the Shapiro-Wilks test were used to assess the normality assumption.

## Results

### Clinical and Demographic Characteristics of the Study Cohorts

The clinical and demographic characteristics of the study population are shown in Tables 1 and 2. Age and racial breakdown were comparable across all three groups (Table 2). Notably, the HFrEF cohort exhibited a male predominance (Table 2). BMI was significantly higher in the HFpEF cohort compared to the other two groups (Table 1). The HFrEF cohort had significantly lower mean systolic and diastolic blood pressure, as well as an elevated mean heart rate (Table 1). Type 2 diabetes hypertension, hyperlipidemia, prior MI, CKD, and atrial fibrillation were significantly higher in the HF groups compared to controls (Table 2). Atrial fibrillation and Type 2 diabetes were both higher in HFpEF versus HFrEF (Table 2).

**Table 1.** Clinical Characteristics of the Study Population.

|  | **Control** | **HFpEF** | **HFrEF** | **Control**<br>**vs. HFpEF**<br>**p-value** | **Control**<br>**vs. HFrEF**<br>**p-value** | **HFpEF**<br>**vs. HFrEF**<br>**p-value** |
| --- | --- | --- | --- | --- | --- | --- |
| **n** | 211 | 357 | 219 |  | n/a |  |
| **Age (Mean(SD))** | 63.8(12.7) | 65.2(11.5) | 62.9(13.4) | 0.3502 | 0.7684 | 0.0758 |
| **Height (in)** | 67.4(3.8) | 67.1(4.5) | 68.6(4.2) | 0.7578 | **0.019** | **0.0007** |
| **Weight (lbs)** | 187(39.3) | 216.5(56.9) | 201.3(55.8) | **<0.0001** | **0.0216** | **0.0056** |
| **BMI (kg/m2)** | 28.9(5.5) | 34.1(8.1) | 29.7(7.1) | **<0.0001** | 0.5266 | **<0.0001** |
| **Systolic BP**<br>**(mmHg)** | 131.6(19.9) | 130.4(19.6) | 117.2(20.8) | 0.79 | **<.0001** | **<.0001** |
| **Diastolic BP**<br>**(mmHg)** | 76(10.9) | 72.4(12.4) | 69.4(15) | **0.0042** | **<.0001** | **0.0192** |
| **Heart Rate**<br>**(BPM)** | 73(12.7) | 77(15.8) | 80.1(16.1) | **0.0089** | **<.0001** | 0.0925 |
| **HBA1C (%)** | 6.2(1.1) | 6.6(1.5) | 6.3(1.3) | 0.068 | 0.8754 | 0.2019 |
| **Serum Glucose**<br>**(mg/dL)** | 103.6(32.6) | 121(48.4) | 116.9(48.8) | **0.0001** | **0.0158** | 0.697 |
| **Creatinine**<br>**(mg/dL)** | .99(.48) | 1.4(1.2) | 1.7(1.6) | **0.0008** | **<.0001** | **0.0395** |

**Table 2.** Demographics and Comorbidities of the Study Population.

|  |  | **Control** | **HFpEF** | **HFrEF** | **Control**<br>**vs. HFpEF**<br>**p-value** | **Control**<br>**vs. HFrEF**<br>**p-value** | **HFpEF**<br>**vs. HFrEF**<br>**p-value** |
| --- | --- | --- | --- | --- | --- | --- | --- |
| **Race** | **White** | 134(63.5%) | 222(62.2%) | 123(56.6%) | 0.69 | 0.29 | 0.15 |
|  | **Black** | 69(32.7%) | 125(35%) | 82(37.4%) |  |  |  |
|  | **Asian** | 1(.5%) | 3(.84%) | 4(1.8%) |  |  |  |
|  | **Other** | 7(3.3%) | 7(1.96%) | 10(4.6%) |  |  |  |
| **Sex** | **Male** | 120(56.9%) | 185(51.8%) | 155(70.8%) | 0.26 | **0.004** | **<.0001** |
|  | **Female** | 91(43.1%) | 172(48.2%) | 64(29.2%) |  |  |  |
| **Type 2 Diabetes (%)** |  | 33(15.6%) | 163(45.7%) | 75(34.2%) | **<.0001** | **<.0001** | **0.0069** |
| **Hypertension (%)** |  | 103(48.8%) | 283(79.2%) | 161(73.5%) | **<.0001** | **<.0001** | 0.11 |
| **Hyperlipidemia (%)** |  | 99(46.9%) | 260(72.8%) | 157(71.7%) | **<.0001** | **<.0001** | 0.76 |
| **Atrial Fibrillation (%)** |  | 16(7.6%) | 202(56.6%) | 100(45.7%) | **<.0001** | **<.0001** | **0.011** |
| **Chronic Kidney Disease (%)** |  | 13(6.2%) | 100(28%) | 77(35.2%) | **<.0001** | **<.0001** | 0.07 |
| **COPD (%)** |  | 16(7.6%) | 100(28%) | 40(18.3%) | **<.0001** | **0.001** | **0.008** |
| **Any Cancer (%)** |  | 6(2.8%) | 98(27.5%) | 49(22.4%) | **<.0001** | **<.0001** | 0.2 |
| **Prior MI (%)** |  | 4(1.9%) | 44(12.3%) | 50(22.8%) | **<.0001** | **<.0001** | **0.001** |

### Fatty Acid Metabolism: Medium- and Long-chain ACs

ACs, intermediates of FAO, were grouped into short-chain (C2-C6, SCAC), MCAC (C8-C12), LCAC (C14-C18), and very long-chain (C20-C22, VLCAC) species. In alignment with prior studies,^18,20,21,23,24^ the majority of unsaturated MCAC and LCAC exhibited elevation in the HFrEF group (Figure 1). These include C8:1-OH, C12-OH, C14:1, C14:1-OH, C14:2, C14-OH, C16:1, C16:1-OH, C16:2-OH, C18:1, C18:1-OH, C18:2, C20:1, and C20:2. Conversely, the saturated MCAC/LCAC/VLCAC species (C8, C10, C12, C14, C16, C18, C20) were not different across the groups (Figure 1). Few ACs were changed in HFpEF.

**Figure 1.**
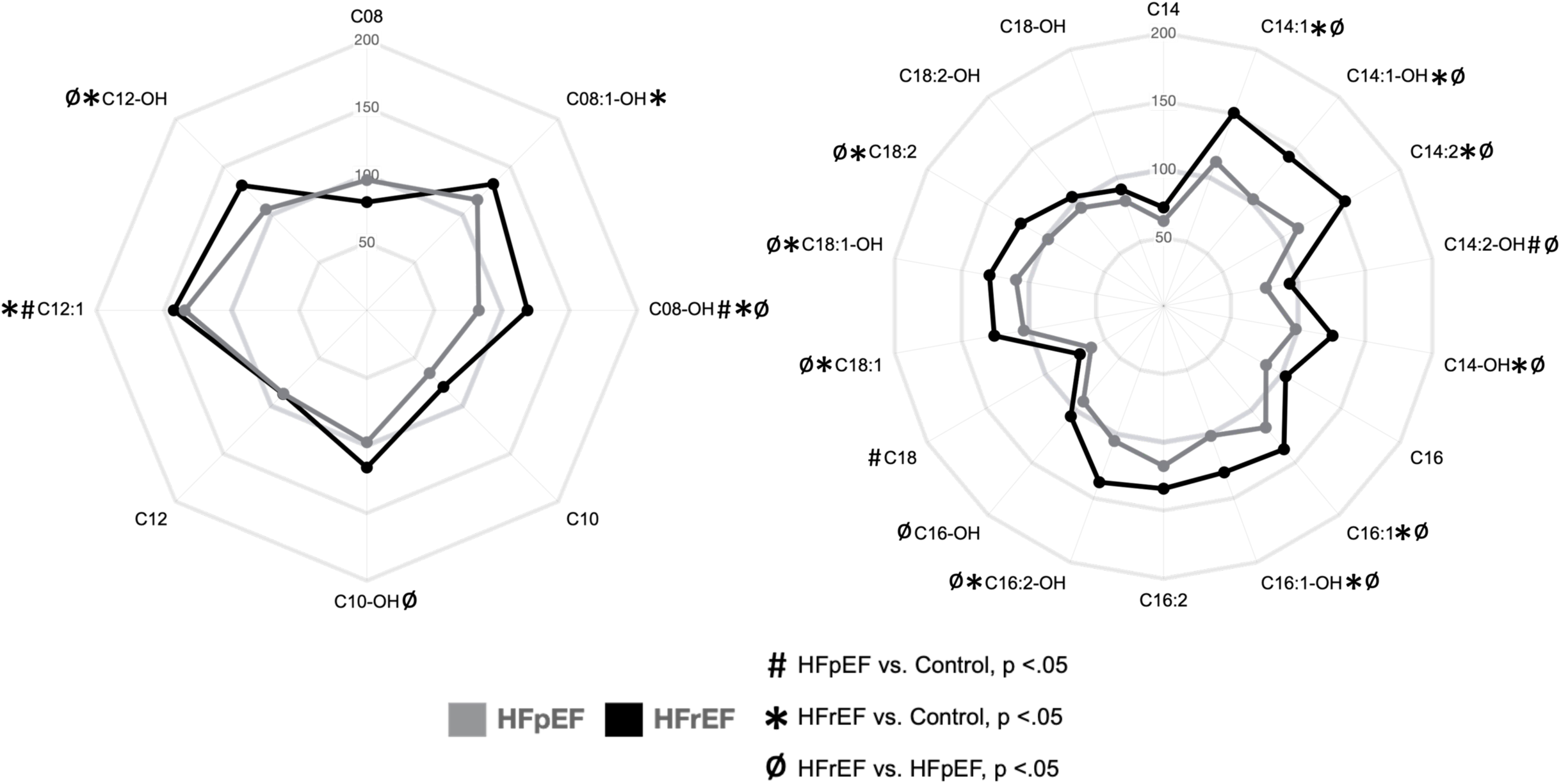
Unsaturated MCAC and LCAC species are elevated in HFrEF but not HFpEF. Radar plots of MCACs (left) and LCACs (right) depicting plasma levels of acylcarnitines in HFpEF (grey) and HFrEF (black) as percentages of control. Symbols indicate significance of comparisons across groups.

### Amino Acid Metabolism: Amino Acids and Amino Acid Derivatives

BCAAs were not elevated in the HF groups in contrast to other reports^14,20,21^ (Figure 2). Rather, levels of leucine were reduced. Notably, however, a number of SCACs that are generated via BCAA degradation were elevated in the plasma of both HF groups (Figure 2). For example, C04-DC methylmalonyl carnitine and C04-DC succinyl carnitine, common downstream metabolites of the degradation of all three BCAAs, were elevated in the plasma of both HF groups against control. C04 isobutyryl carnitine demonstrated a similar pattern. C04-OH isobutyryl, C05-2-methylbutyryl, C05:1, C05-OH, and C06-OH carnitines exhibited stepwise elevation, highest in HFrEF (Figure 2).

**Figure 2.**
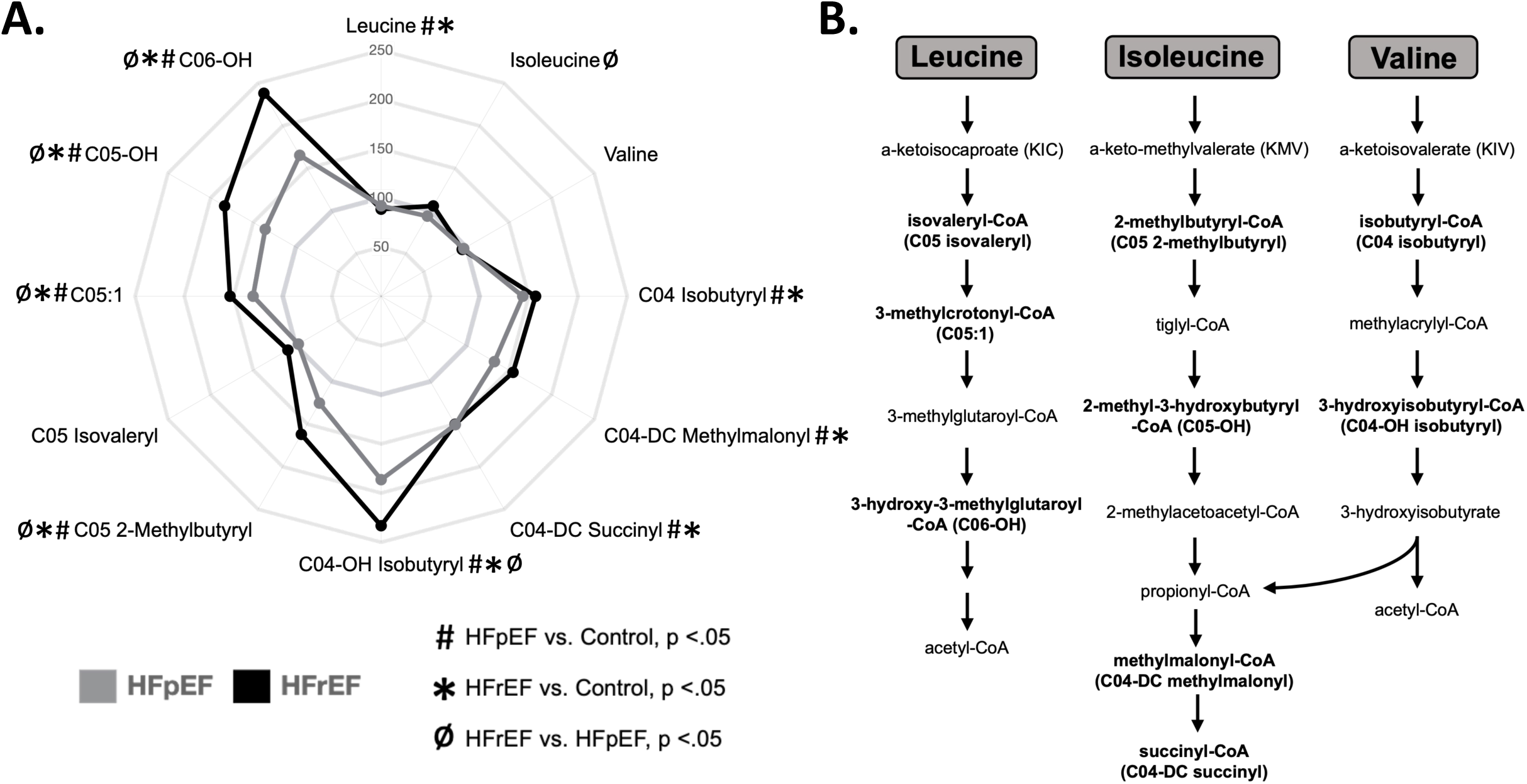
Plasma metabolome in both HFrEF and HFpEF subjects reflect increased BCAA degradation. **A)** Radar plot depicting plasma levels of branched-chain amino acids (BCAA) and BCAA metabolites in HFpEF (grey) and HFrEF (black) as percentages of control. Symbols indicate significance of comparisons across groups. **B)** Diagram displaying the BCAA metabolic pathway. Metabolites in bold represent acylcarnitine species measured in this study. SCAC correlates are indicated in parenthesis.

A number of amino acid metabolites exhibited notable changes in the HF groups. 1-methylhistidine demonstrated a stepwise increase across the HF groups, with highest levels in the HFrEF cohort (Figure 3). 3-methylhistidine was similarly elevated in the two HF groups compared to control. Two arginine derivatives known to regulate the biosynthesis of nitric oxide were also increased in the HF groups: asymmetric dimethylarginine (ADMA) and symmetric dimethylarginine (SDMA) (Figure 4A). SDMA was significantly elevated in both HF groups whereas ADMA, a direct inhibitor of nitric oxide synthase,^30^ was uniquely elevated in HFpEF. Arginine was unchanged across the groups (Figure 4B). In addition, mean plasma homocitrulline levels were increased in both HF groups, and were more than doubled in the HFrEF group over control (Figure 4D). Citrulline, kynurenine, phenylalanine, and tyrosine were also elevated in both forms of HF compared to control (Figure 3).

**Figure 3.**
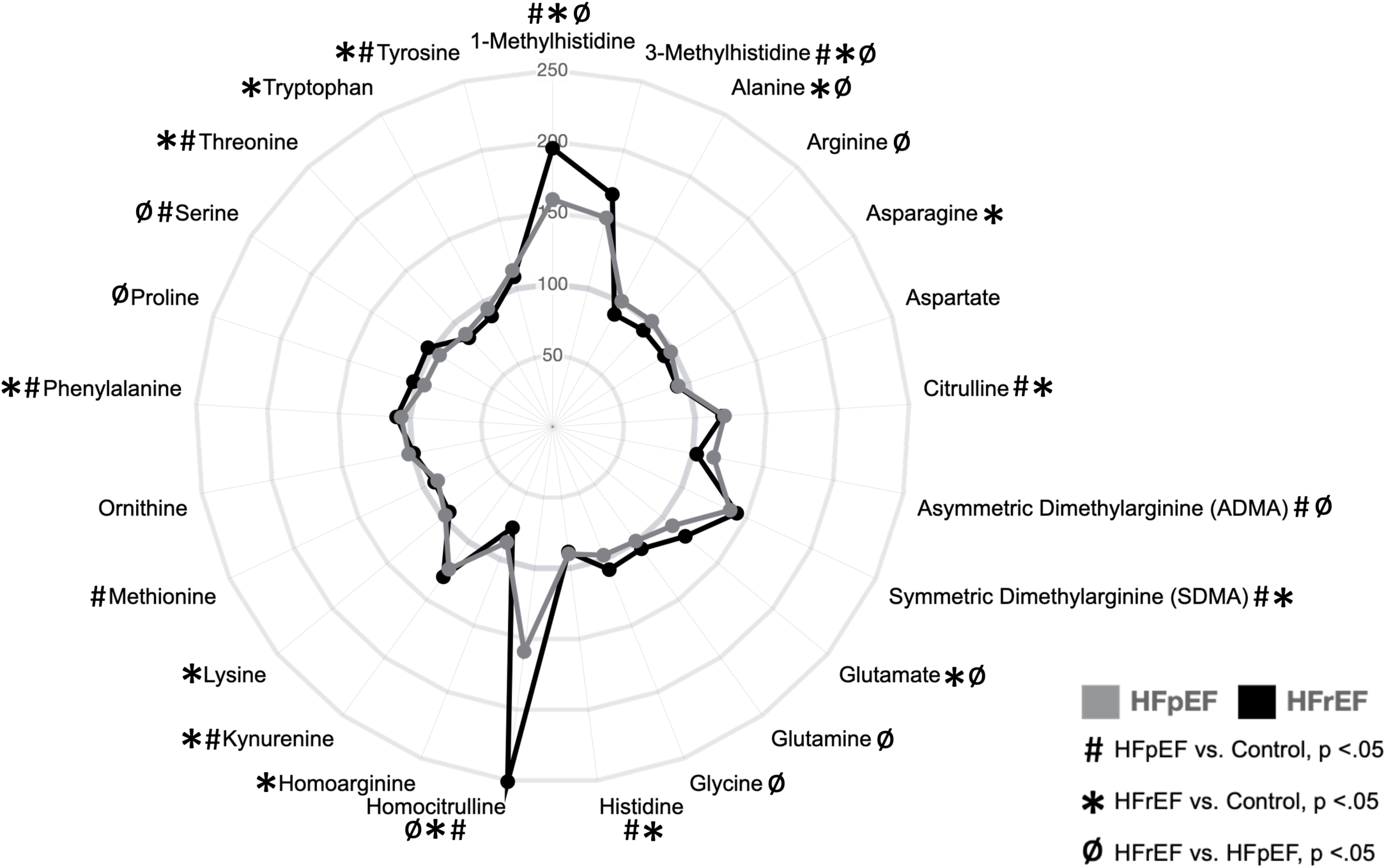
Levels of a subset of amino acids are elevated in the plasma of HF patients. Radar plot depicting plasma levels of amino acids in HFpEF (grey) and HFrEF (black) as percentages of control. Symbols indicate significance of comparisons across groups. Notably elevated species included 1- and 3-methylhistidine, symmetric dimethylarginine (SDMA), asymmetric dimethylarginine (ADMA), and homocitrulline.

**Figure 4.**
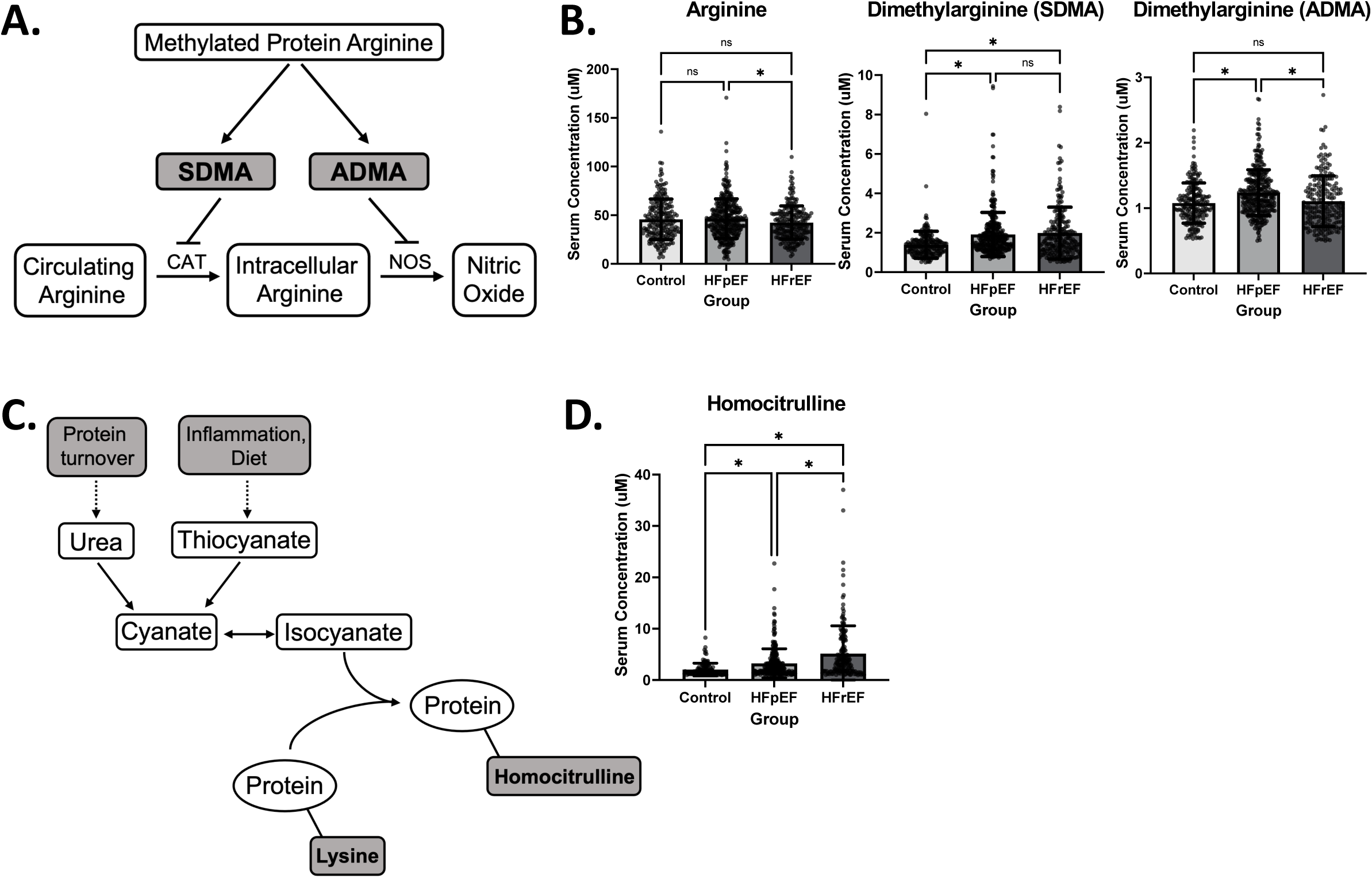
Asymmetric dimethylarginine is uniquely elevated in HFpEF while homocitrulline is elevated in both forms of HF. **A)** Metabolic pathway depicting the formation of asymmetric dimethylarginine (ADMA) and symmetric dimethylarginine (SDMA) and subsequently their effects on arginine metabolism. **B)** Bar graphs depicting mean +/- SD of serum concentrations of Arginine (left), SDMA (middle), and ADMA (right) by cohort. Asterisk indicates p-value < .05. **C)** Metabolic pathway depicting the formation of homocitrulline residues from the incorporation of isocyanate into proteins with lysine residues. **D)** Bar graph depicting mean +/- SD of serum concentrations of Homocitrulline by cohort, showing significant elevation in HFrEF. Asterisk indicates p-value < .05.

### Tricarboxylic Acid Cycle (TCA) and Ketone Metabolism: Organic Acids

TCA metabolites citrate, succinate, fumarate, and malate were mildly but significantly elevated in the plasma of both HF groups (Figure 5A). The most notable elevations in the organic acid category were observed for alpha-ketoglutarate and 3-HBA, which were both uniquely elevated in HFrEF. In addition, C4-OH carnitine, a metabolite downstream of 3-HBA oxidation, was similarly uniquely elevated in HFrEF (Figure 5B).

**Figure 5.**
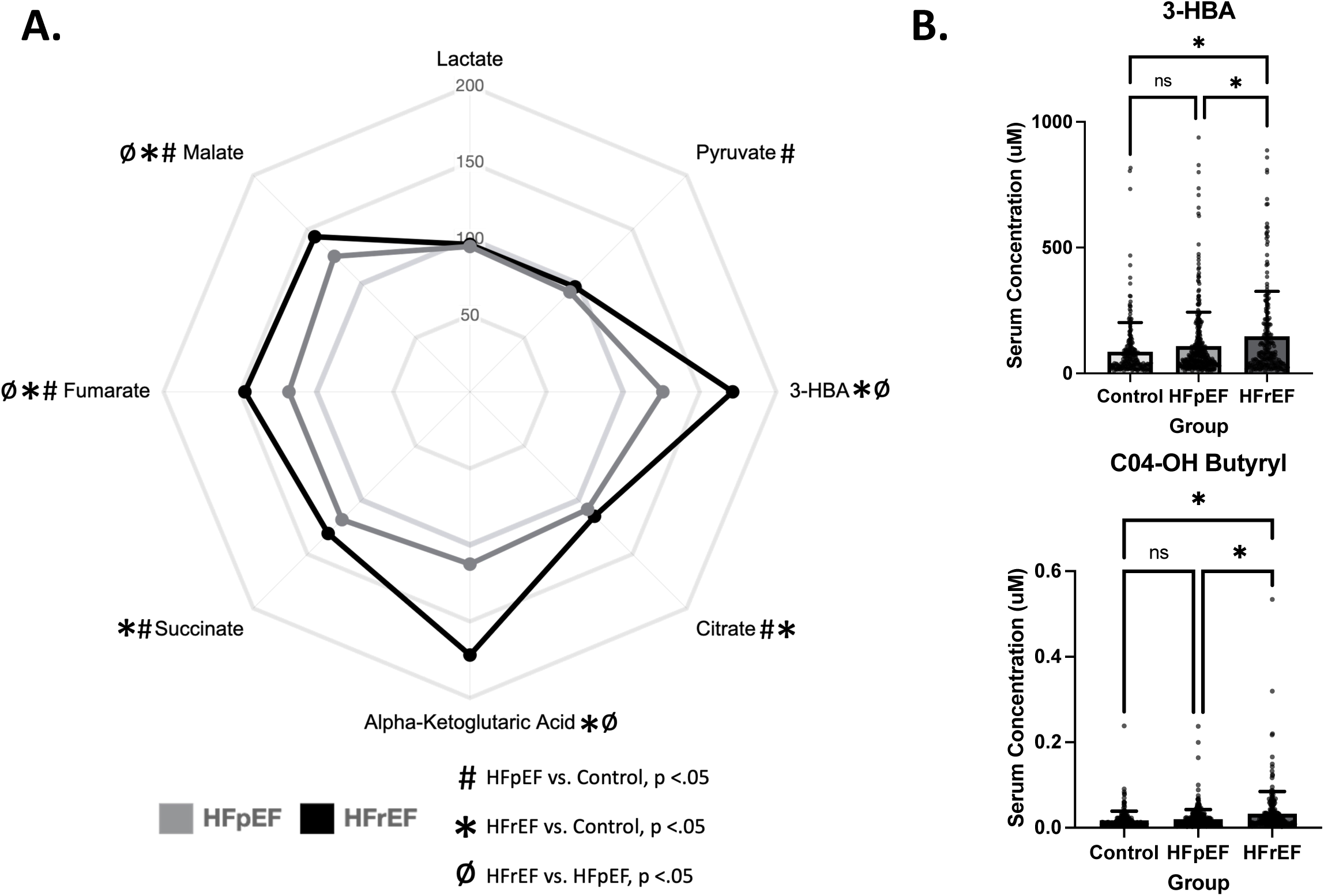
Plasma levels of ketone body 3-hydroxybutyrate (3-HBA), and its downstream metabolite, C4-OH carnitine, are uniquely elevated in HFrEF subjects. **A)** Radar plot depicting plasma levels of organic acids in HFpEF (grey) and HFrEF (black) as percentages of control. Symbols indicate significance of comparisons across groups. **B)** Bar graphs depicting mean +/- SD of serum concentrations 3-HBA (top) and its metabolite C4-OH carnitine (bottom), which are both uniquely elevated in HFrEF. Asterisks indicate p-value <.05.

### Correlation of NT-proBNP with 3-HBA and C4-OH Carnitine

The observation that 3-HBA is uniquely elevated in the HFrEF group strongly suggests that hepatic ketogenesis is increased in this group. In addition, elevation of C4-OH carnitine in the HFrEF group is a marker of increased ketone body oxidation. An increase in circulating ketone bodies has been described in HFrEF patients^5,16,18,31,32^ but the underlying mechanism that links HF to increased hepatic ketogenesis is unknown. Ketosis often occurs in the context of increased free FA delivery to the liver, secondary to adipose lipolysis. Previous studies have indeed shown increased plasma free fatty acid (FFA) levels in patients with HFrEF.^33–35^ Given that natriuretic peptides are known to increase lipolysis, we hypothesized that BNP-mediated lipolysis may lead to increased hepatic ketogenesis in the HFrEF group.^36^ NT-proBNP levels were elevated in both forms of HF compared to controls with significantly higher levels in the HFrEF group (Figure 6A). Notably, hypertension and diabetes had no significant impact on NT-proBNP levels. In linear regression models, a positive correlation between 3-HBA and NT-proBNP was observed in both HFrEF and HFpEF, with the strongest association in the HFrEF cohort (Figure 6B). Notably, plasma 3-HBA and NT-proBNP were not significantly correlated in the control group. A similar pattern was seen for the ketone metabolite C4-OH carnitine (Figure S1). Taken together these results suggest a potential mechanism whereby increased natriuretic levels generated by the failing heart results in lipolysis and increased delivery of FFA to the liver, which in turn increases FAO and ketogenesis (Figure 6C).

**Figure 6.**
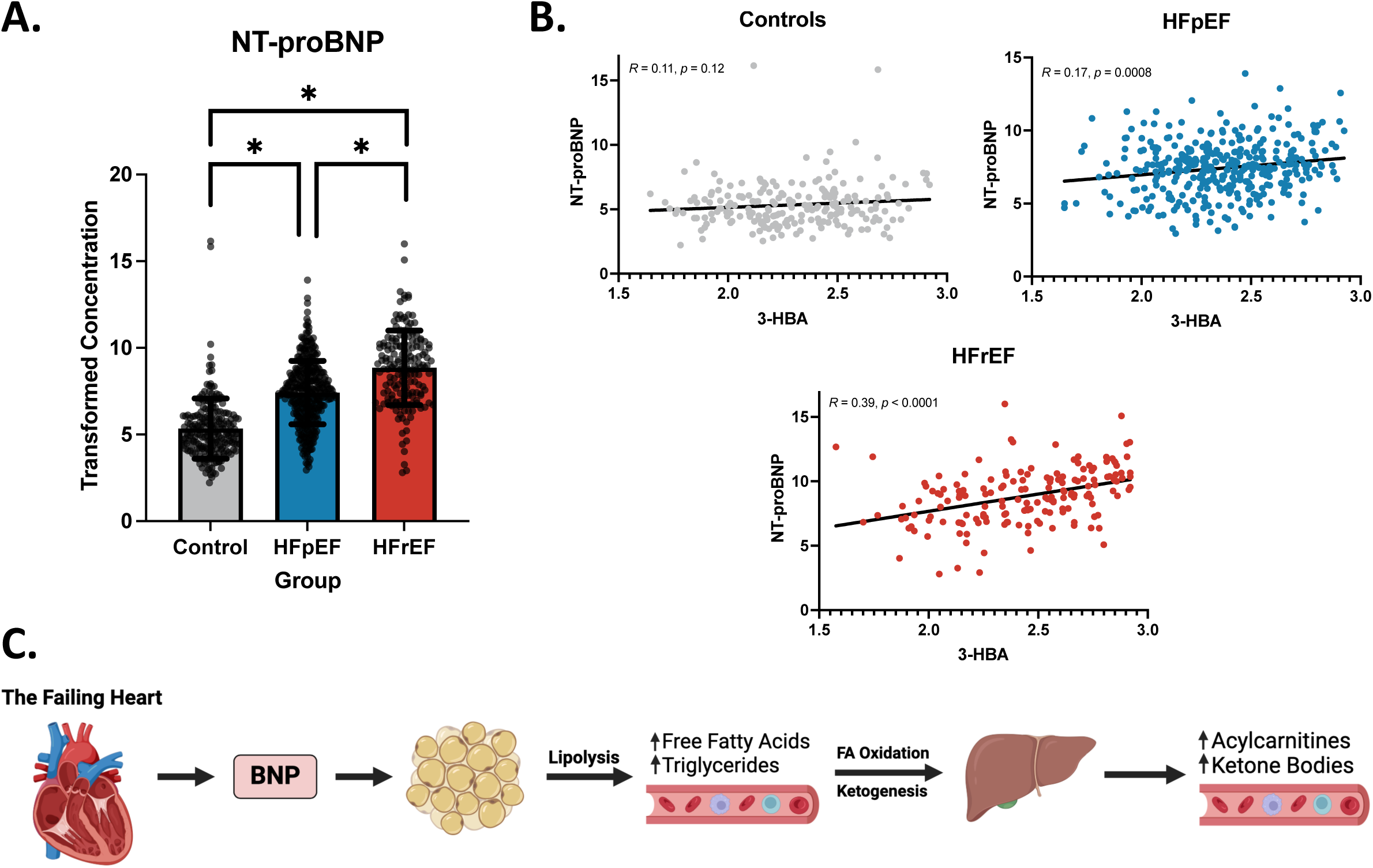
NT-proBNP is significantly elevated in HFrEF and is correlated with levels of 3-HBA in patients with HF but not in the Control group. **A)** Bar graphs depicting mean +/- SD of transformed concentration of plasma NT-proBNP in controls, HFpEF, and HFrEF. Asterisks indicate p-value <.05. **B)** Linear regressions depicting the relationship between the plasma concentration of 3-HBA and the transformed plasma concentration of NT-proBNP within control (top left), HFpEF (top right), and HFrEF (bottom). **C)** Proposed mechanism by which BNP indirectly acts to increase serum acylcarnitines and ketone bodies. In response to volume overload and increasing wall stress, the failing heart secretes natriuretic peptides, including BNP. BNP is known to act on adipocytes to increase lipolysis, leading to increased serum FFA and triglycerides. Subsequently, the liver oxidizes the FAs generating increased levels of acylcarnitine intermediates and acetyl-CoA, the latter serving as a substrate for ketone synthesis. Created with BioRender.com.

## Discussion

This study identifies several novel findings and reinforces previous findings regarding comparative plasma metabolite levels in patients with HFrEF, HFpEF, and controls without HF in a relatively large and diverse population using targeted quantitative metabolomics. The samples were collected in a manner in which time of last meal was not controlled, enhancing the potential for application to biomarker discovery and development. Our results confirmed the results of previous studies as well as identified new metabolite markers of HF, including both shared and HF phenotype-specific signatures. First, as has been described, we found that circulating MCACs, LCACs, and the ketone body 3-HBA were increased in patients with HFrEF compared to HFpEF and controls.^18,20,21,23,24,31^ Second, in contrast to a few published studies,^14,20^ BCAA levels were not different among the groups but metabolites of BCAA degradation were increased in both HFrEF and HFpEF. Third, our results identify a number of interesting novel amino acid metabolite signatures including elevation of methylated amino acid derivatives in both HF groups. Furthermore, a methylated arginine derivative, asymmetric dimethylarginine, was increased uniquely in the HFpEF group. Lastly, we find that the levels of circulating NT-proBNP correlated with 3-HBA and C4-OH carnitine across the heart failure groups, suggesting a potential axis between the failing heart, adipose tissue, and the liver.

This study corroborates several previous studies that have found LCACs to be elevated in the plasma of patients with HF. More specifically, we found that a large subset of unsaturated MCAC and LCACs were uniquely elevated in HFrEF. Very few of these species were significantly changed from control in HFpEF. Our findings are most similar to those of Hunter et al.^20^ and Hahn et al,^18^ in which ACs were found to be elevated in HF, with highest levels in HFrEF compared to HFpEF. Furthermore, two independent studies investigating the plasma of patients with ischemic HF and diminished EF, patients that would mostly be described in the current HF schema as HFrEF, found that the HF groups had significantly elevated ACs across various chain lengths.^23,24^ On the other hand, some studies have reported elevation of AC species in HFpEF greater than HFrEF.^21,22^ A unique pattern in our acylcarnitine profile was the consistency with which saturated species were unchanged in either form of HF. This pattern has only been reported once before in a small subset of patients with end-stage dilated cardiomyopathy.^17^ The basis for this observation is unclear but may reflect the composition of the triglyceride pool in the adipose tissue which in turn is influenced by diet. It is also possible that the activity of FA desaturases is reduced in some tissues in HFrEF.

While several preclinical models of HF have shown increased levels of circulating BCAAs,^14,37^ human plasma metabolomic analyses have failed to consistently corroborate this finding. Hunter et al. found that BCAAs and related metabolites were uniquely elevated in the plasma of patients with HF.^20^ Furthermore, in a combined preclinical and clinical study, Sun et al. found an increase in the circulating branched chain keto acid α-keto-β-methylvalerate.^14^ However, our results, as well as those of Hahn et al., did not find a significant change in circulating BCAAs in humans with HF.^18^ In fact, we found a modest downregulation of leucine in the HF groups. The reason for these varying findings remains unclear, but inconsistent dietary restriction and fasting status across the studies is likely a contributing factor. Furthermore, it is possible that other study populations had a greater prevalence of diabetes or insulin resistance in the HF cohorts compared to controls which could influence the results. Interestingly, we found a pattern of increased SCACs indicative of catabolism of BCAAs in the HF groups (Figure 2). This was also seen to some degree by Hahn et al.^18^ This is a surprising finding given that transcriptomic and metabolomic studies of failing myocardium suggest reduced BCAA degradation in HF.^3,14,37^ Accordingly, the source of the increased BCAA oxidation is likely extra-cardiac such as skeletal muscle or liver. Notably, pharmacologic or genetic interventions that increase BCAA catabolism have been shown to improve or attenuate HF in mouse models of HF.^14,37,38^ Future studies will be needed to further investigate the origin of these metabolites and what role they may play, compensatory or pathologic, in HF.

The panel of amino acid metabolites analyzed uncovered a number of interesting and novel findings. One particularly noteworthy finding was an elevation of ADMA, an arginine metabolite, uniquely in HFpEF. Its counterpart, SDMA, was elevated in both forms of HF. These species are known to interfere with nitric oxide synthesis by either directly or indirectly inhibiting nitric oxide synthase (NOS) (Figure 4A), contributing to worsening systemic vasoconstriction.^30^ These species have also been found to be positively correlated with NT-proBNP and with increased severity of diastolic dysfunction in patients with HFpEF.^39^ Whether they are involved as a driver of HF or merely a metabolic consequence of HF remains to be determined. Increased systemic vasoconstriction and cardiac afterload via NOS inhibition could contribute to pathologic myocardial hypertrophy that ultimately manifests as HF. Furthermore, coronary vasomotor disturbances could also be contributory.^39^ Notably, increasing emphasis has been placed on the potential role of vascular dysregulation as a driver of HFpEF.^40^ Further studies will be needed to determine whether ADMA and SDMA can be used as biosignatures for earlier identification of HFpEF and whether they can be utilized as targets for vascular modulation to ameliorate or prevent HF.

Homocitrulline was elevated in a stepwise fashion in the HF groups with the HFrEF group exhibiting over twice the mean plasma concentration of the control group. L-homocitrulline is an amino acid and a metabolite of ornithine metabolism. It is believed that the depletion of the ornithine supply causes the accumulation of carbamyl-phosphate in the urea cycle which may be responsible for the enhanced synthesis of homocitrulline and homoarginine.^41^ It is important to note that the homocitrulline measured in metabolomics assays is likely a combination of diet-based factors as well as a measure of protein carbamylation, which is the incorporation of cyanate residues into amino acids secondary to abundance of serum urea.^42^ While homocitrulline has not been extensively studied in HF, it has been positively correlated with NT-proBNP as well as broadly implicated in cardiovascular disease, chronic kidney disease, atherosclerosis, and endothelial dysfunction.^42,43^ We also found that 1- and 3-methylhistidine were elevated in the HF groups over control. These species have not been previously studied in HF. They are thought to be biomarkers of skeletal muscle protein turnover and toxicity.^44^ Thus, they may be useful prognostic biomarkers that allow clinicians to determine the severity and decompensation of patients with HF.

Studies in humans have shown that HF triggers an increase in circulating ketone bodies.^5,16,18,21,31,45^ In addition, evidence from our group and others ranging from pre-clinical studies to humans found that the failing heart shifts to utilizing ketone bodies as a compensatory ancillary fuel.^5,16,19,46^ Our results are consistent with this, showing an increase in 3-HBA, also known as beta-hydroxybutyrate (BHB), and C4-OH carnitine in the HFrEF group. Hahn et al. similarly found increased ketogenesis in HFrEF as opposed to HFpEF.^18^ However, other studies have found either no change in HF versus control^20^ or have found that ketones are increased in HFpEF to a greater degree than HFrEF.^21^ The mechanism whereby HF triggers hepatic ketogenesis in HF is unknown. Furthermore, it is unclear as to why 3-HBA levels are higher in HFrEF relative to HFpEF as observed in this study and by others. Natriuretic peptides are leading candidates for metabolic signaling from the failing heart. BNP is known to mediate lipolysis,^36^ and our results found that NT-proBNP was higher in HFrEF compared to HFpEF, as has been described previously.^47^ Indeed we found a significant correlation between levels of NT-proBNP and 3-HBA and its downstream metabolite C4-OH carnitine in our HF cohorts. We hypothesize that the failing heart secretes BNP which ultimately drives increased lipolysis resulting in increased FFA delivery to the liver, which in turn increases FA oxidation (and increased production and secretion of MCACs and LCACs) and generates increased levels of acetyl-CoA, a substrate for ketogenesis (Figure 6C). This is most predominant in HFrEF given that NT-proBNP was most significantly upregulated in this group. Further studies will be needed in order to assess the degree to which ketosis in HF is BNP-dependent and whether this pathway can be harnessed for new therapeutic avenues.

### Study Limitations

In spite of the robust sample size, racial diversity, and wide metabolomic panel, this study has several limitations. Notably, by not controlling fasting status and using a singular plasma sample, our results are susceptible to day-to-day dietary variability. This is likely limited by the large sample size, however it remains a factor to consider. Additionally, the collection of plasma samples at routine outpatient visits ensures that the majority of the HF cohorts are likely well-compensated, which does not accurately reflect the totality of the HF population. Our controls were matched to the HFpEF group for comorbidities and demographics, which could possibly exaggerate the findings in the HFrEF group. Lastly, while we analyzed an extensive plasma metabolomic panel, these findings are provided without the context of corresponding myocardial tissue analysis, which ultimately would have contributed to the interpretation of our findings.

## Data Availability

The authors confirm that the data supporting the findings of this study are available within the article and/or its supplementary materials.

## Acknowledgements

Metabolomics studies were performed by the Metabolomics Core (RRID:SCR_022381) in the Penn Cardiovascular Institute and supported, in part, by NCI P30 CA016520 and NIH P30 DK050306-27. We thank Stephanie DerOhannessian for help with sample collection and preparation. We acknowledge the Penn Medicine BioBank (PMBB) for providing data and thank the patient-participants of Penn Medicine who consented to participate in this research program. The PMBB is approved under IRB protocol# 813913 and supported by Perelman School of Medicine at University of Pennsylvania, a gift from the Smilow family, and the National Center for Advancing Translational Sciences of the National Institutes of Health under CTSA award number UL1TR001878.

## Non-standard Abbreviations and Acronyms

HF: heart failure
HFrEF: heart failure with reduced ejection fraction
HFpEF: heart failure with preserved ejection fraction
FAO: fatty acid oxidation
FA: fatty acid
FFA: free fatty acid
AC: acylcarnitine
SCAC: short-chain acylcarnitines
MCAC: medium-chain acylcarnitines
LCAC: long-chain acylcarnitines
BCAA: branched-chain amino acids
3-HBA: 3-hydroxybutyrate
BHB: beta-hydroxybutyrate
ADMA: asymmetric dimethylarginine
SDMA: symmetric dimethylarginine
BNP: brain/b-type natriuretic peptide
NT-proBNP: N-terminal pro b-type natriuretic peptide
Nitric Oxide Synthase: NOS
aKG: alpha-ketoglutarate

